# Immunotherapy Time of Infusion Impacts Survival in Head and Neck Cancer: A Propensity Score Matched Analysis

**DOI:** 10.1101/2024.01.08.24300992

**Authors:** Daniel A. Ruiz-Torres, Saskia Naegele, Archana Podury, Lori Wirth, Sophia Z. Shalhout, Daniel L. Faden

## Abstract

The adaptive immune response is physiologically regulated by the circadian rhythm. Data in lung and melanoma malignancies suggests immunotherapy infusions earlier in the day may be associated with improved response; however, the optimal time of administration for patients with HNSCC is not known. We aimed to evaluate the association of immunotherapy infusion time with overall survival (OS) and progression free survival (PFS) in patients with HNSCC in an Institutional Review Board-approved, retrospective cohort study. 113 patients met study inclusion criteria and 98 patients were included in a propensity score-matched cohort. In the full unmatched cohort (N=113), each additional 20% of infusions received after 1500h conferred an OS hazard ratio (HR) of 1.35 (95% C.I.1.2-1.6; p-value=0.0003) and a PFS HR of 1.34 (95% C.I.1.2-1.6; p-value <0.0001). A propensity score-matched analysis of patients who did or did not receive ≥ 20% of infusions after 1500h showed that those who were administered ≥20% of infusions after 1500h trended towards a shorter OS (HR=1.35; p-value=0.26) and a shorter PFS (HR=1.57, 95% C.I. 1.02-2.42, p-value=0.04). Each additional 20% of infusions received after 1500h remained robust in the matched cohort multivariable analysis and was associated with shorter OS (adjusted HR=1.4 (95% C.I.1.2-1.8), p-value<0.001). Patients with advanced HNSCC who received more of their infusions in the afternoon were associated with shorter OS and PFS and scheduling immunotherapy infusions earlier in the day may be warranted.

**Conflict of Interest Statement:** Dr. Wirth reports receiving advisory board fees from Ayala Pharmaceuticals, Blueprint Medicines, Cue Biopharma, Cullinan Oncology, Genentech USA, Loxo Oncology, Merck, NewLink Genetics, Novartis, and Rakuten Medical, consulting fees and advisory board fees from Bayer HealthCare Pharmaceuticals and Eisai, advisory board fees and fees for serving on a steering committee from Eli Lilly, and fees for serving on a data and safety monitoring board from Iovance Biotherapeutics. Dr. Faden has received research funding from Bristol Myers Squibb and Foundation Medicine, holds equity in Illumina and receives consulting fees from Noetic and Focus on Boston. The remaining authors have no conflicts to report.

**Research Highlights:**

- Immunotherapy early in the day may result in improved response rates in HNSCC, consistent with data in other solid malignancies.

## INTRODUCTION

Immunotherapy has emerged as the standard of care in patients with recurrent and/or metastatic head and neck squamous cell carcinoma (HNSCC). However, most HNSCC patients progress on immunotherapy with a response rate of approximately 15-20%.^1-4^ With such heterogenous outcomes, paired with high costs, strategies to optimize the efficacy of immunotherapy and facilitate the adaptive immune system’s anti-tumor response are needed.

An increasing body of evidence suggests dependence of the immune response on the circadian rhythm, which provides organisms the ability to properly predict and respond to environmental cyclical changes and oscillations.^5-9^ The intrinsic molecular clocks of immune cells play an essential role in influencing the innate and adaptive immune response pathways. For example, a randomized trial demonstrated a greater antibody response in subjects upon administration of the influenza vaccine in the morning.^10,11^ Notably, the reproducible cyclical recruitment of immune cells has implications for both cancer and cancer treatment, for example, by guiding tumor infiltrating T-cells. Emerging studies suggest that cancer treatments are influenced by the circadian rhythm.^12-16^ For example, a randomized controlled trial demonstrated improved efficacy with the chronomodulation of oxaliplatin-based chemotherapy for colorectal cancer versus a constant-rate infusion method.^17^ A phase III trial supported the potential for sex to influence chronotherapy and demonstrated improved overall survival in male colorectal cancer patients but not females, in a study that chronomodulated the infusion of fluorouracil, leucovorin, and oxaliplatin versus conventional delivery.^18-20^ Chronomodulation of immune checkpoint inhibitors has also recently been investigated in melanoma and non-small cell lung cancer (NSCL), demonstrating improved survival in patients administered infusions in the morning compared to the afternoon.^21-23^ In keeping with this emerging field of study, we aimed to assess if the specific time of immunotherapy infusions might impact survival outcomes in HNSCC patients.

## METHODS

### Study cohort and outcomes

We performed a cohort study approved by the Mass General Brigham Institutional Review Board. The Mass Eye and Ear and Massachusetts General Hospital Cancer Data Registries were used to retrospectively identify patients with recurrent, advanced or metastatic HNSCC that were treated with immune checkpoint inhibitors (ICI; pembrolizumab, nivolumab, ipilimumab, durvalumab or any combination of these ICIs or chemoimmunotherapy) from January 1, 2016, to June 30, 2022. Clinicopathologic features were extracted from the electronic health records and data was de-identified via the Safe Harbor Method. Immunosuppression was defined as any patient with non-indolent hematological malignancies including chronic lymphocytic leukemia, non-Hodgkin lymphoma, and multiple myeloma, any solid organ or stem cell transplant recipients or patients with a long-standing prior history of immunosuppressive therapy.

Overall survival (OS) was indexed from the date of initiation of ICI to the date of death from any cause or censored at date of the last follow-up visit before database lock of March 31, 2023, in the cases without a death event. Progression-free survival (PFS) was indexed from the date of initiation of ICI therapy to the earliest date of progressive disease (PD) or death or censored at date of the last follow-up visit before database lock in the cases without PD or death events. The primary outcome assessed the association between the proportion of infusions received after 1500h and OS and PFS.

### Statistical analysis and propensity score matched analysis

The proportion of infusions of ICI received after 1500h for every patient was assessed for association with OS and PFS using Cox proportional hazard regression and propensity score-matching in an exploratory analysis. 1500h was designated as the cutoff for afternoon/evening based on several factors including the closing time of the institutional administering immunotherapy infusion clinics and guided by previous definitions of afternoon/evening cutoffs in chronomodulation/chronotherapy studies. Importantly, we determined *a priori* that the afternoon timepoint cutoff to be used in this analysis would be denoted based on the hour that allowed for the maximum number of patients in the propensity score-matched cohort to allow for a well-powered analysis. This was determined by comparing total number of patients available in 30-minute increments starting from 1400h. For example, 1500h maximized the number of patients for propensity score matching at 98 compared to only 22 patients total at a cutoff of 1630h. The proportion of infusions of ICI received after the cutoff was partitioned into increments of twenty percent for each patient and hazard ratios (HR) of OS and PFS were determined for increasing quintiles.

The propensity score-matched cohort was determined by splitting the full unmatched cohort into two infusion groups- those who received at least 20% of their ICI infusion after cutoff and those who did not-using logistic regression and a width of 0.1 caliper via backward selection for final variable selection: stage, prior surgery, prior radiotherapy, prior systemic therapy, HPV status and immunosuppression.^24^ A proportion of 20% was used to maintain consistency with previous ICI chronotherapy studies assessed in melanoma and NSCLC patients for comparison. This still allowed for ideal comparison groups in our cohort.^21-23^ To assess if the matched cohort was well-balanced in terms of features, clinicopathological characteristics between the two propensity score-matched infusion groups were assessed using χ^2^ test for categorical variables and the Kruskal-Wallis test for continuous variables.

OS and PFS were assessed with Kaplan-Meier method and reported with 95% confidence intervals (C.I.). The estimated PFS HR was adjusted in multivariable Cox proportional hazards regression for the matched patients with the propensity score matched analysis variables and the estimated OS HR was adjusted in a backward selected multivariable Cox proportional hazards regression on the matched cohort. The statistical tests performed were two-sided and the reported p-values were exploratory in nature only. Therefore, no Bonferroni corrections were applied, and reported associations are centered on the overall totality of evidence where predictors were observed to maintain trends in the univariable and multivariable models. Statistical analyses were performed in R (version 4.2.2).

## RESULTS

The full cohort that met inclusion criteria included 113 patients with recurrent or advanced stage HNSCC who received immunotherapy treatment with a median follow-up time of 1.1 years [IQR 0.8-1.7] (**Figure 1**). In this complete unmatched cohort, the median age was 65 years (range: 28-96) at the start of ICI. Seventy-eight (69%) patients were male, and 53 (47%) had a smoking history ≥10 Pack-Year. One hundred patients (88%) had an Eastern Cooperative Oncology Group (ECOG) performance status ≤ 1. The majority of the cohort (95 patients; 84%) received pembrolizumab, were administered ≤ 6 infusions of immunotherapy (64 patients; 57%), and 7 (6%) developed adverse events that led to the complete discontinuation of ICI therapy. The primary cause of any treatment cessation was progressive disease. (**Table 1**) Twenty-seven (24%) patients had HPV-associated HNSCC, and 5 (4%) were immunosuppressed. Eighty-one patients (72%) received prior chemotherapy (**Supplemental Table1**), 84 (74%) received prior surgery and 105 (93%) patients received radiotherapy for HNSCC. Thirteen patients (14%) also received cytotoxic chemotherapy in combination with ICI. Additional clinicopathological characteristics are described in Table 1.

**Figure 1.**
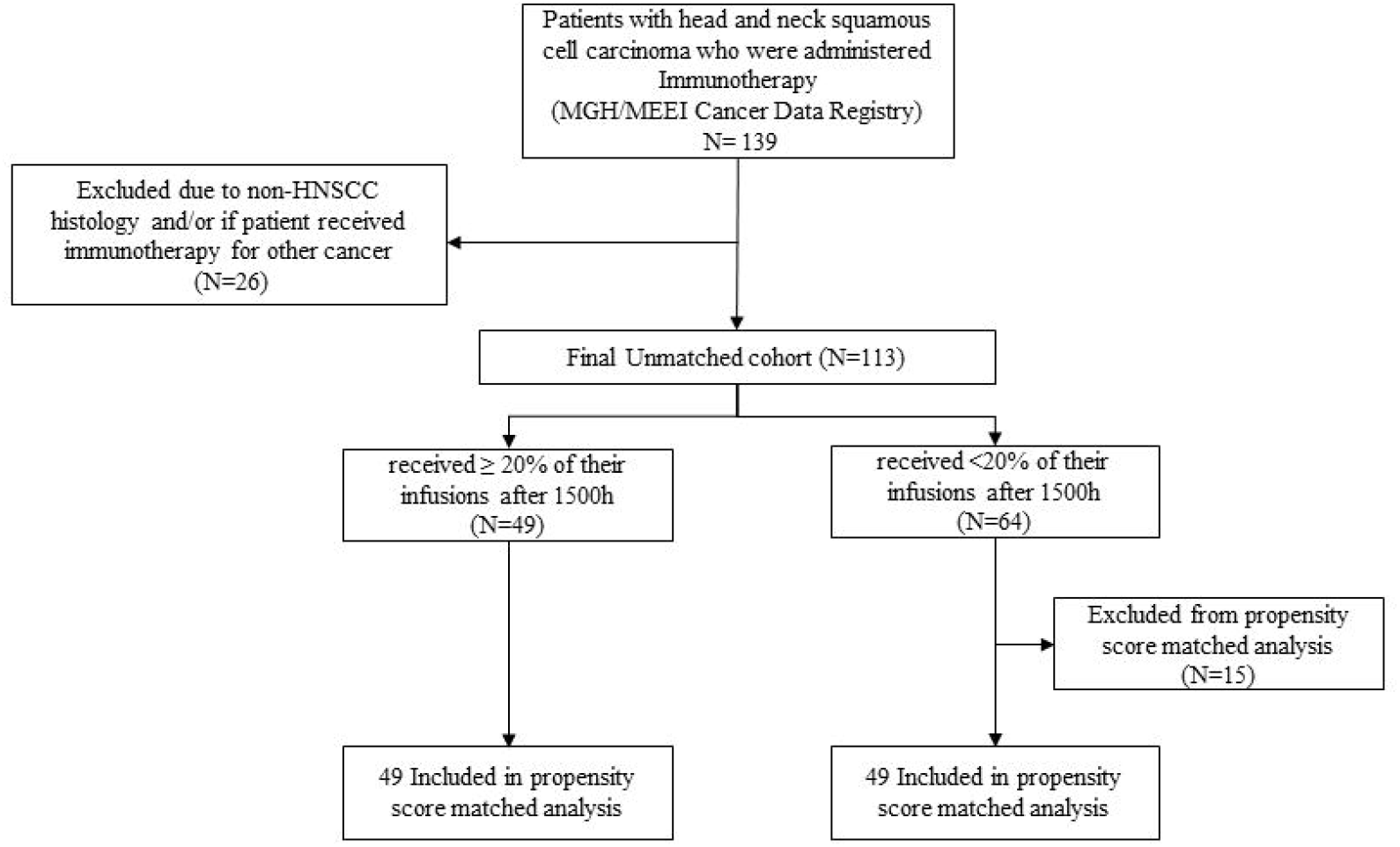
Consort Diagram/Study Profile: Study overview of patients at Mass Eye and Ear and Mass General Cancer Center that received immunotherapy between January 2016 to June 2022 for advanced HNSCC. The propensity score matching variables: stage, receipt of radiotherapy, prior surgery, prior chemotherapy, immunosuppression and HPV-status.

Forty-nine patients (43%) received at least 20% of ICI infusions after 1500h (**Figure 1**). Each additional twenty percent of ICI infusions received after 1500h conferred an OS HR of 1.35 (95% C.I. 1.2-1.6; p-value= 0.0003) and a PFS HR of 1.34 (95% C.I. 1.2-1.6; p-value <0.0001).

Upon propensity score matching, the 49 patients (43%) who received at least 20% of ICI infusions after 1500h were matched with 49 patients who did not. Clinicopathological characteristics were well-balanced between the two infusion groups (**Table 1**). The median age in each cohort infusion group was similar (65 years (range:44-92) versus 63 years (range:28-96), p-value=0.86), as was ECOG ≤ 1 (41 patients (84%) versus 45 patients (92%), p-value= 0.47) and smoking history ≥10 Pack-Year (26 patients (53%) versus 22 (45%), p-value=0.42). In this study, there were no ICI treatment related deaths. Adverse events that led to the complete cessation of ICI were balanced between the two comparison infusion groups (2 cases (4%) versus 5 cases (10%), p-value=0.44). Of note, there appears to be more males in the group of patients who received < 20% of infusions after 1500h. However, on univariable analysis of the matched cohort, sex was not associated with OS (HR= 0.81, 95% C.I. 0.46-1.45, p-value=0.49) or PFS (HR= 0.74, 95% C.I. 0.46-1.19, p-value=0.21) (**Supplemental Table2**). Furthermore, type/primary site of HNSCC, and previous therapy including surgery, radiation and/or chemotherapy, were not associated with OS or PFS in the matched cohort analysis (**Supplemental Table 2**). The total number of infusions appears higher in the group of patients who received <20% of infusions after 1500h. As previously described in recent studies assessing ICI chronomodulation in NSCLC and melanoma, number of infusions is a variable that is directly dependent on the outcome. Often, the number of total infusions is determined by the managing clinicians directly based on the patient’s durable response to ICI, or lack thereof, and toxicity.^21,22^ In this study, more infusions received were associated with an OS advantage (HR= 0.11, 95% C.I. 0.05-0.21, p-value < 0.001) and longer PFS (HR= 0.09, 95% C.I. 0.05-0.17, p-value < 0.001) in the matched cohort, as one may expect.^21,22^

Receiving at least twenty percent of ICI infusions after 1500h trended towards a shorter OS (HR= 1.35, 95% C.I. 0.81-2.25, p-value=0.26) in the propensity score matched analysis and full unmatched cohort study (HR=1.48, 95% C. I. 0.92-2.38, p-value=0.1). (**Figure 2**) Receiving at least twenty percent of ICI infusions after 1500h was associated with a shorter PFS (HR= 1.57, 95% C.I. 1.02-2.42, p-value=0.04) in the propensity score matched analysis and remained robust to multivariable analysis when adjusted for stage, prior surgery, radiation, chemotherapy and HPV status (adjusted HR=1.58, 95% C.I. 1.02-2.46, p-value= 0.04). (**Table 2**) Receiving at least twenty percent of ICI infusions after 1500h was also associated with a shorter PFS in the full unmatched cohort study (HR=1.67, 95% C. I. 1.11-2.51, p-value=0.01). (**Figure 2**). Finally, each additional 20% of infusions received after 1500h remained robust in the matched cohort multivariable analysis and was associated with shorter OS (adjusted HR=1.4 (95% C.I. 1.2-1.8), p-value< 0.001) (**Figure 3**).

**Figure 2.**
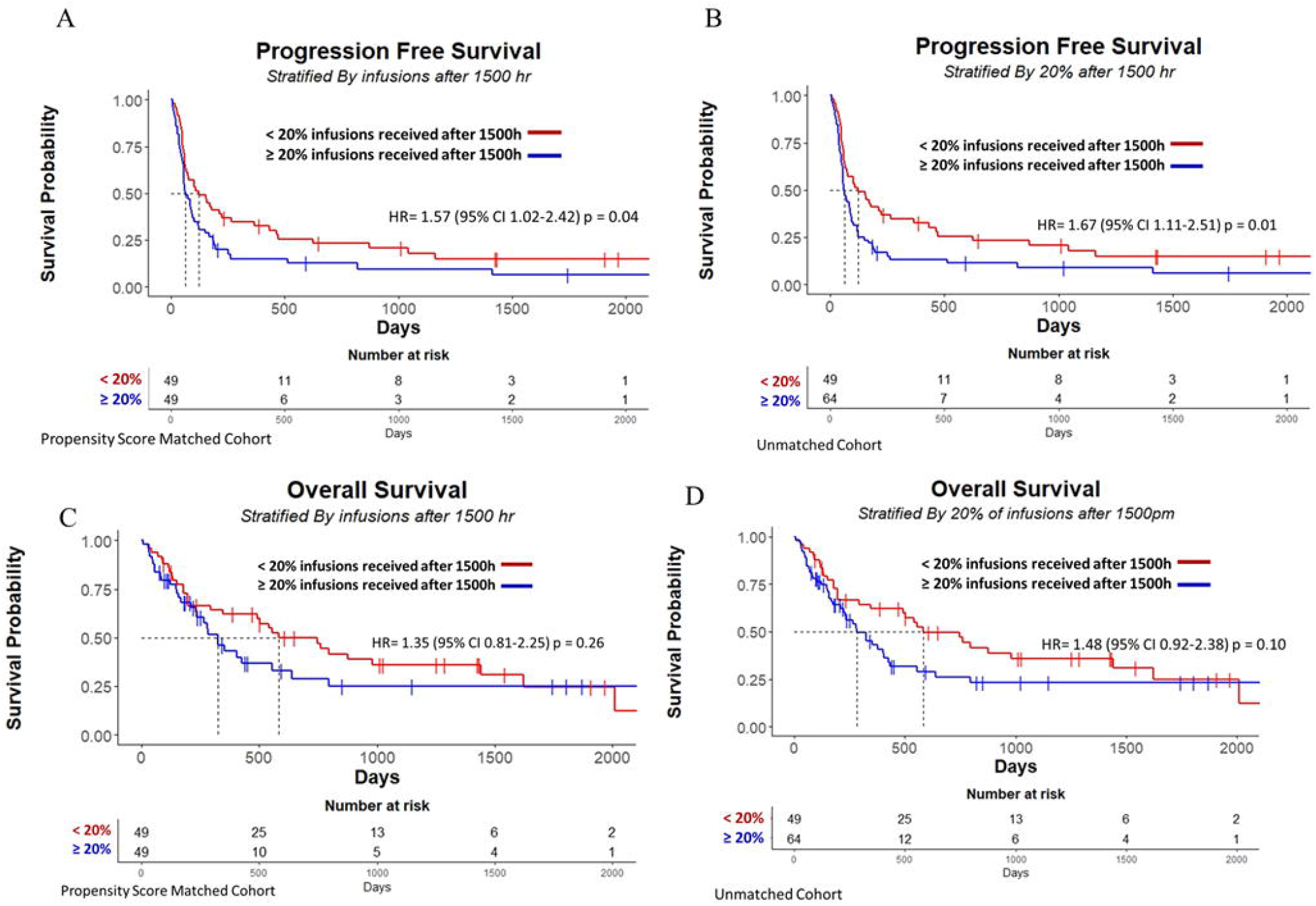
Kaplan Meier Curve of Progression Free Survival and Overall Survival: Progression Free Survival for propensity score-matched groups (A) and unmatched groups (B). Overall Survival for propensity score-matched groups (C) and unmatched groups (D).

**Figure 3.**
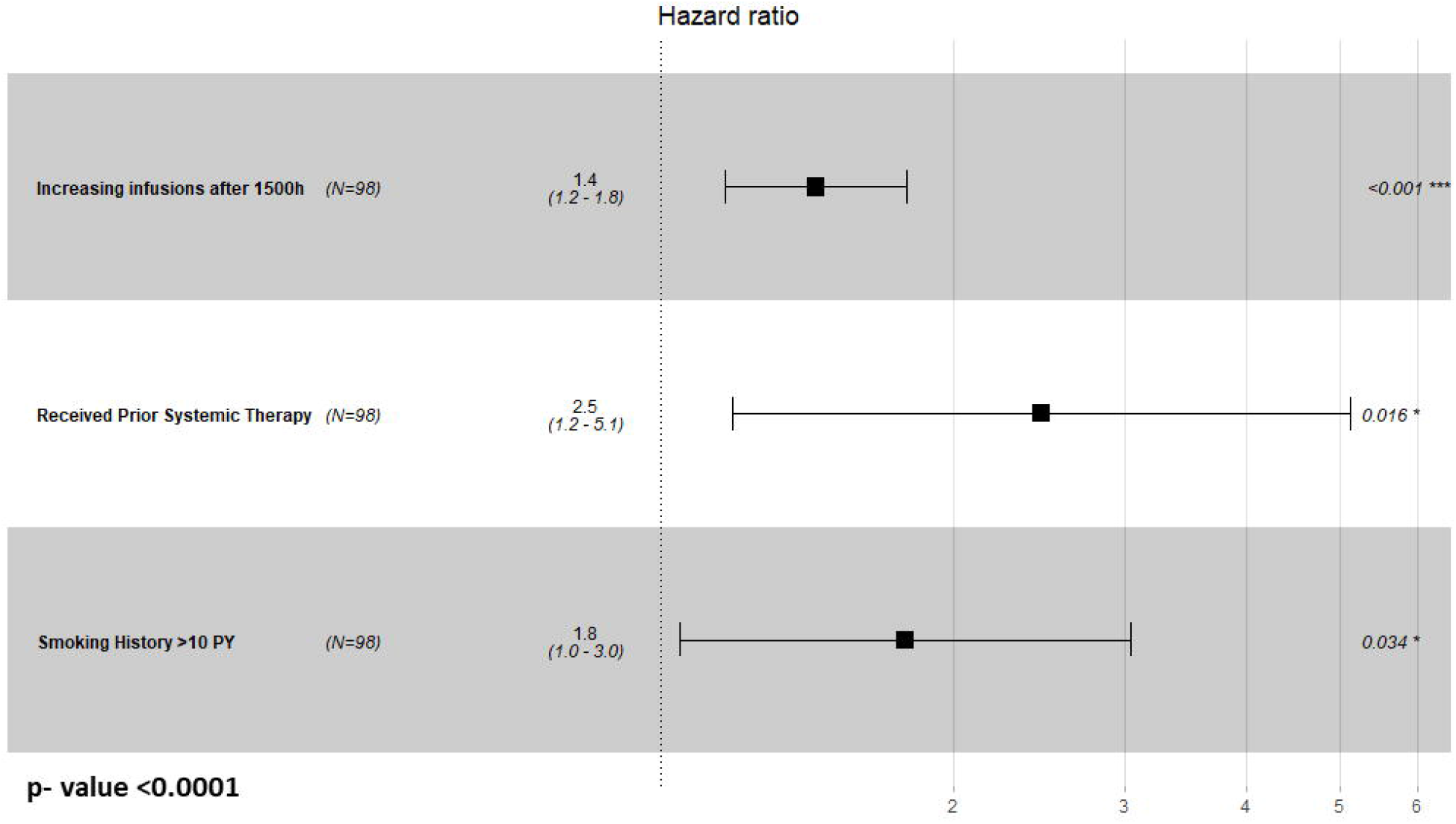
Forest Plot of Overall Survival: Multivariable Cox proportional hazards regression of overall survival, in the propensity score-matched groups of patients who did (n=49) and did not (n=49) receive ≥20% of infusions after 1500h.

## DISCUSSION

In our retrospective cohort study of patients with advanced HNSCC, receiving an increasing number of infusions in the afternoon was observed to be associated with a worse OS and PFS in the full unmatched cohort and remained robust in a multivariable analysis of the propensity score matched cohort. Receiving at least 20% of ICI infusions after 1500h was associated with a worse PFS in the full unmatched cohort analysis, the propensity score-matched analysis and remained robust in a multivariable Cox proportional hazards regression model. Furthermore, receiving at least 20% of ICI infusions after 1500h also trended to a worse OS in the matched and unmatched cohorts of patients with HNSCC.

While previous studies have assessed chronomodulation of cancer treatment including interferon, cytotoxic chemotherapy, and interleukin 2, few studies have directly assessed the impact of ICI timing on OS and PFS. To our knowledge, no previous studies have assessed ICI chronotherapy for patients with HNSCC.^17,25-27^ In a recent retrospective trial that assessed time-of-day patterns of ICI infusions in advanced Stage IV melanoma patients, a strong association was observed with those receiving at least 20% of ICI infusions after 1630h and a worse OS.^21^ A pilot study that assessed time-of-day patterns in patients with NSCLC treated with ICI found that those who received nivolumab in the morning were associated with a better overall response and those patients with primary resistance to nivolumab were most often observed in the patients receiving afternoon infusions. ^22,23^ However, these studies excluded patients who received less than 4 doses which is a previously highlighted and thoroughly discussed caveat of those analyses.^22^ For example, some patients continue to experience durable response to 4 or less doses of ICI despite cessation of therapy due to an adverse event, or receive 4 or less doses primarily due to progressive disease.^28^ These cases still warrant study of a potential chronobiological influence especially since recent analyses suggest the first few doses of ICI may be the most important for driving response or toxicity in melanoma.^21,29^ In this context, the inclusion of these patients may impact the evaluation of chronotherapy of ICI.^21,22^ We chose to include such patients to determine if chronomodulation of ICI reports remain robust in the absence of this major selection bias since most patients with primary resistance to ICI receive less than 4 doses.^22^ The HNSCC cohort assessed in this study primarily received ≤ 6 doses and therefore exclusion of ≤ 4 doses would not be representative of real-world practices of ICI in this disease setting and substantially limit study sample size (**Table 1**). Nonetheless, the results of our study support the findings of these previous studies, despite inclusion of these cases.

While we did perform a propensity score-matched analysis to achieve a balance between prior treatment history, stage, and HPV-status in the two comparison infusion groups, a major limitation of our study is the retrospective nature. Further, studies have shown differences in the circadian rhythm of males versus females, but the majority of our cohort was male.^19,20^ Male predominance is typical for a HNSCC cohort and on univariable analysis, sex had no impact on OS and PFS and was not found to be significant in the multivariable analysis and therefore removed from the final model due to step-wise backward selection. However, a more representative cohort may be necessary to elucidate sex differences in ICI chronomodulation and chronotherapy. Additionally, a larger study that enrolls patients from a wider variation in locations and latitudes, since natural light exposure influences the circadian rhythm, would also allow for more generalizable findings.

Despite the heterogenous cohort used in this analysis, age, HPV-status, site of HNSCC, previous therapy, and stage were not associated with OS and PFS on univariable analysis (**Supplement Table 2)**. The association of infusions after 1500h remained robust upon multivariable analysis accounting for these and other confounding variables (**Table 1**) and therefore these features were removed from the final multivariable model due to step-wise backward selection. (**Figure 3**).

Finally, the total number of infusions of ICI received by a patient may have an impact or contribute bias that would be more appropriately quantified in a randomized controlled prospective trial. In this retrospective study, patients who received a greater number of ICI infusions were associated with a longer OS and PFS. However, we have excluded the total number of infusions as a predictor or covariable in both the propensity score-matched analysis and multivariable analysis because total number of infusions actually serves as a surrogate for survival and/or response and does not appropriately meet the conditions for a confounding variable, as previously described in ICI chronomodulation studies.^21,22^ Despite these caveats, our study suggests that prioritizing infusions of immunotherapy earlier in the day for patients with HNSCC may extend both PFS and OS by aligning therapy administration with the patient’s optimal adaptive immune response capabilities as regulated by the circadian rhythm. With only an ~20% response rate for patients with HNSCC, and high drug costs, optimization of ICI therapy is critical.

This work supports continued investigation focused on the circadian clock’s dynamic 24h cycle and its effect on the immune system, tumor infiltrating lymphocytes and T-cells. Furthermore, time-of-day pattern studies may also improve responses to immunotherapy beyond ICI, including adoptive T-cell therapy and oncolytic viral therapy. For example, circulating T-cells were observed at higher rates in healthy subjects in the morning compared to the evening with the defined circadian regulation of trafficking of lymphocytes delineated in previous studies.^30,31^ The circadian clock’s influence on T-cell and B-cell trafficking further support aligning such therapies with the 24h established cycle.^32-35^

## CONCLUSIONS

In-keeping with other recent studies, administering immunotherapy earlier in the day may confer improved progression free survival and overall survival, further supporting that aligning immunotherapy treatment to exploit the regulation of the adaptive immune response via the circadian rhythm may offer clinical benefit. Considering these findings, and those in other solid malignancies, a prospective multi-site trial is warranted to further investigate timing of immunotherapy infusions. Furthermore, studies assessing the mechanisms underlying ICI induced stimulation of the immune response and dependencies on the circadian clock to optimize treatment management for HNSCC patients are also needed.

## Data Availability

All data produced in the present study are available upon reasonable request to the authors

