## Supplemental Table 1 and 2 for "Immunotherapy Time of Infusion Impacts Survival in Head and Neck Cancer: A Propensity Score Matched Analysis"

**Specialty Topic: Immunotherapy/Systemic Therapy**

**Authors:** Daniel A. Ruiz-Torres, MD^1^, Saskia Naegele, MD^1^, Archana Podury, MD^1^, Lori Wirth^3^, Sophia Z. Shalhout, PhD^1^**^#*^**, Daniel L. Faden, MD^1,2^**^#*^**

**Affiliations:**

^1^Department of Otolaryngology-Head and Neck Surgery, Massachusetts Eye and Ear, Harvard Medical School, Boston, MA 02114 USA

^2^ Broad Institute, Cambridge MA USA

^3^ Massachusetts General Hospital Cancer Center, Massachusetts General Hospital, Boston, MA 02114 USA.

***These authors contributed equally and are joint senior authors.**

**#Corresponding Authors:**

Sophia Shalhout, PhD

Department of Otolaryngology-Head and Neck Surgery

Massachusetts Eye and Ear, Harvard Medical School,

Boston, MA 02114 USA

Daniel L. Faden, MD

Department of Otolaryngology-Head and Neck Surgery

Massachusetts Eye and Ear, Harvard Medical School,

Boston, MA 02114 USA

**Supplementary Table 1:** Details for Prior Systemic Therapy

| Supplementary Table 1:  Details for Prior Systemic Therapy (N = 113) | |
| --- | --- |
| Patient Characteristics | **Full Cohort N=113**  **No. (%)** |
| Prior Systemic Therapy* | **63 (56)** |
| ≥3 chemotherapeutic agents | 16 (14) |
| 2 chemotherapeutic agents | 15 (13) |
| 1 chemotherapeutic agent | 32 (28) |

*Chemotherapy agents included alkylating agents, anti-metabolites, anti-mitotic drugs and EGFR inhibitors.

**Supplementary Table 2:** Univariable Analysis for Overall Survival (OS) and Progression Free Survival (PFS) in Matched Cohort

| Supplementary Table 2: Univariable Analysis for OS and PFS | | |
| --- | --- | --- |
|  | **Matched Cohort (N=98)** | |
| Patient Characteristics | OS HR (95% C.I.) p-value) | PFS HR (95% C.I.) p-value |
| Age | 1.0 (0.98-1.03)   p-value=0.62 | 1.0 (0.98-1.02)   p-value=0.87 |
| Sex | 0.81 (0.46-1.45)  p-value=0.49 | 0.74 (0.46-1.19)  p-value=0.21 |
| Race | 3.52 (1.33-9.34)  p-value=0.01 | 2.03 (0.97-4.3)  p-value=0.06 |
| Smoking History ≥10 Pack-Year | 1.63 (0.98-2.76)  p-value=0.06 | 1.62 (1.05-2.50)  p-value=0.03 |
| ECOG Performance status | 1.64 (1.07-2.52)  p-value=0.02 | 1.08 (0.75-1.55)  p-value=0.67 |
| Primary site | 0.90 (0.73-1.11)  p-value=0.32 | 0.98 (0.82-1.17)  p-value=0.83 |
| Stage | 2.6 (0.36-18.92)  p-value=0.34 | 0.96 (0.34-2.63)  p-value=0.93 |
| Anti-PD-1 therapy | 1.08 (0.51-2.31)  p-value = 0.60 | 2.25 (1.17-4.32)  p-value=0.10 |
| HPV Status | 0.95 (0.67-1.34)  p-value=0.76 | 0.87 (0.65-1.17)  p-value=0.37 |
| Number of Infusions | 0.11 (0.05-0.21)  p-value < 0.001 | 0.09 (0.05-0.17)  p-value < 0.001 |
| Season of first infusion | 1.47 (0.71-3.07)  p-value=0.50 | 1.06 (0.57-1.98)  p-value=0.80 |
| Received prior systemic therapy | 1.55 (.78-3.06)  p-value=0.211 | 1.49 (0.88-2.52)  p-value=0.13 |
| Received prior surgery | 1.36 (0.77-2.43)  p-value=0.29 | 1.04 (0.65-1.68)  p-value=0.85 |
| Radiotherapy | 0.54 (0.19-1.51)  p-value=0.30 | 0.51 (0.22-1.19)  p-value=0.20 |
| Increasing infusions after 1500h | 1.32 (1.09-1.61)  p-value=0.004 | 1.34 (1.12-1.6)  p-value=0.001 |
